# State AIDS Drug Assistance Programs’ Contribution to the United States’ Viral Suppression, 2015-2022

**DOI:** 10.1101/2025.04.04.25325288

**Authors:** Kathleen A. McManus, Amy Killelea, Erin Q. Rogers, Feng Liu, Tim Horn, Amber Steen, Jessica Keim-Malpass, Auntré Hamp, Elizabeth T. Rogawski McQuade

## Abstract

**Background:** State AIDS Drug Assistance Programs (ADAPs) provide HIV medication access for people with HIV (PWH) with low incomes in the United States (US). We quantified the proportion of viral suppression (VS) that is from ADAP clients for 2015-2022.

**Methods:** For 2015-2022, we obtained viral load (VL) test results and VS data from publicly available, jurisdiction-level data on ADAP clients and PWH. We report descriptive statistics including the proportion of PWH with a VL who had VS and were supported by ADAPs.

**Results:** After excluding jurisdictions with missing data, PWH who were included in the analysis for each year was 63.7-96.4%. VS for PWH each year was 60-66.3%. VS for ADAP clients was 81.2%-91.4%. In all years, compared to all PWH, a lower proportion of ADAP clients had a reported VL and a higher proportion had VS. Over 2015-2022, the average proportion of PWH who were ADAP clients was 23.1%, the proportion of PWH with VLs who were ADAP clients was 22.2%, and the proportion of PWH with VS who were ADAP clients was 30.8%.

**Conclusions:** Almost a third of the entire VS rate was from ADAP clients, despite ADAP serving less than 25% of PWH. A much higher proportion of ADAP clients achieved VS, compared to PWH. ADAPs’ impact is not due to ADAP clients being over represented among PWH with reported VLs. ADAP does not directly receive any federal Ending the HIV Epidemic (EHE) Initiative funding. Policymakers should examine how ADAPs can support the EHE Initiative.

**Summary:** Despite state AIDS Drug Assistance Programs (ADAPs) serving less than a quarter of people with HIV, almost a third of the viral suppression in the United States can be attributed to ADAPs. ADAPs are essential for ending the HIV epidemic.

## Introduction

AIDS Drug Assistance Programs (ADAPs) are a key component of the Ryan White HIV/AIDS Program (RWHAP), which was enacted in 1990 and has been a vital safety net for low-income people with HIV (PWH). ADAPs, which are managed by state health departments, receive federal funding to provide HIV and other essential medications to low-income PWH without insurance. ADAPs also help low-income insured clients to pay for their premiums and HIV-related cost sharing. In 2022, ADAPs served over 235,000 clients or 20% of the 1.1 million PWH (both diagnosed and undiagnosed) in the United States (US).^1^

Each ADAP has a formulary that enumerates the medications the program covers for uninsured clients as well as the medications for which ADAP covers cost sharing for insured clients. By statute, ADAP formularies must include antiretroviral therapies (ART).^2^ As the US Food and Drug Administration approves new ART, ADAPs have expanded their formularies to include new medications, including single tablet and long-acting injectable regimens.^3^ ADAPs have also expanded their formularies to include other HIV-related medications outside of ART, which has been increasingly important to address co-morbidities. ADAPs receive federal funding that allows them to procure medications for their clients. Medications are then distributed via the health department pharmacy or a network of retail pharmacies in the state. ADAP clients often also receive a host of other RWHAP services depending on their income and the services available where they live, including HIV-related clinical care, case management, and support services.

When compared with other peer high-income countries, such as the United Kingdom, France, and Canada, the US has the highest ART list prices and the lowest rate of HIV viral suppression (VS).^4^ Average list prices of ART are over $4,000 per patient per month with annual price increases outpacing overall inflation rates.^5^ While ADAPs are entitled to steep discounts from the list price because of their participation in the federal 340B Drug Pricing Program and their ability to negotiate additional discounts directly from HIV drug manufacturers, the net price of medications still impacts ADAP formulary decisions and client access.^5^ ADAPs insulate PWH from these costs by providing the medication to low-income PWH without insurance and by helping insured clients to pay for their premiums and medication cost sharing.

An essential aspect of the US federal Ending the HIV Epidemic (EHE) plan is to “rapidly and effectively” provide ART to PWH to achieve sustained VS.^6^ VS is a key measure of HIV treatment efficacy and is achieved when PWH have less than 200 copies the virus per milliliter of blood.^5^ Once someone is virally suppressed, they cannot transmit the virus to someone else.^7^ However, ADAPs are not eligible to directly receive any EHE funding. Instead, Phase one EHE funding has been focused on 42 RWHAP Part A recipients (city and county governments in metropolitan areas), seven RWHAP Part B recipients (state health departments), and 32 CDC HIV prevention grantees (state and local health departments).^8^

To date, there have been no longitudinal studies examining the contribution of ADAP to the VS in the US. Such studies are necessary to understand how ADAPs contribute to ending the HIV Epidemic in the US. We quantified the percent of VS in the US that is due to ADAP for 2015 to 2022.

## Methods

### Data sources

In this retrospective longitudinal study, we analyzed aggregated, state-level data from 51 U.S. jurisdictions (50 states and the District of Columbia (DC)) for calendar years 2015 through 2022. Data on ADAP clients were obtained from the National Alliance of State and Territorial AIDS Directors (NASTAD) *National RWHAP Part B and ADAP Monitoring Project Annual Reports*. The data from annual reports for 2017 to 2024 were used for this study.^1,9–13^ For each jurisdiction-year, we collected the number of ADAP clients served, the number of ADAP clients eligible for VS assessment, and the number of ADAP clients with VS. In the NASTAD data, an ADAP client was *eligible* for VS assessment if they had a reported viral load (VL) test during that calendar year. VS was defined as the last reported VL of the year being < 200 copies/ml.^1,9–13^

Corresponding data were obtained for all PWH from the Centers of Disease Control and Prevention’s (CDC) National Center for HIV/AIDS, Viral Hepatitis, STD, and TB Prevention (NCHHSTP) AtlasPlus.^14^ For each jurisdiction-year, we collected the number of PWH, the number of PWH eligible for VS assessment, and the number of PWH with VS. In the CDC data, a person was *eligible* for VS assessment if diagnosed by the end of the previous calendar year and alive at the end of analysis year. VS was defined as the last reported VL of the year being < 200 copies/ml.^15^ We derived measures for PWH who are not ADAP clients by subtracting the ADAP client counts from the total PWH counts in each jurisdiction-year.

The University of Virginia Institutional Review Board for Human Subjects Research determined this study to be non-human subject research.

### Analysis

All 51 jurisdictions were considered each year; a jurisdiction was included in a given year’s analysis if both ADAP and CDC data were complete. We excluded a jurisdiction for that year if either the ADAP or CDC data lacked VL or VS data, or if the CDC data were internally inconsistent (i.e., the reported number eligible for VS calculation exceeded the total number of PWH in that jurisdiction).

For each jurisdiction-year, we calculated descriptive metrics for all PWH, ADAP clients, and non-ADAP clients: the percentage eligible for VS assessment, the percentage who had VS (among those eligible), and the percentage who had a detectable VL (among those eligible). We further quantified ADAP’s contribution to outcomes each year by calculating the proportion of PWH who were ADAP clients, the proportion of PWH who were eligible for VS assessment who were ADAP clients, the proportion of PWH with VS who were ADAP clients, and the proportion of individuals with a detectable VL who were ADAP clients.

Our primary analysis measured the proportion of PWH with VS and proportion of PWH with a detectable VL. We tested the robustness of our findings based on different imputation strategies to assess the impact of missing VL data on key outcomes. Three alternate scenarios were considered for individuals missing a reported VL in a given year: (1) assuming all unknown VLs were detectable (i.e., all missing counts treated as detected VLs, a worst-case scenario); (2) assuming the same VS rate for the unknown VLs as observed among those with reported VLs (base-case scenario) (3) assuming all unknown VLs were virally suppressed (treating all missing as undetectable VLs, a best-case scenario). We performed the same sensitivity analyses for the proportion of PWH with detectable VLs.

We used a chi-square test to compare ADAP clients vs. non-ADAP clients on all observed outcomes, including sensitivity analyses. Recognizing the potential for Type 1 error, we used a Bonferroni-adjusted alpha of 0.0007 (0.05 divided by 72 independent tests). All analyses were performed using R 4.4.2 software (R foundation, Vienna, Austria).

## Results

### Study Population

A total of 51 jurisdictions contributed data over the study period, though some were excluded in certain years due to missing data. The estimated number of PWH in the United State increased from 942,988 in 2015 to 1,092,023 in 2022. After applying inclusion criteria, the percent of PWH in the US who were included in the study for each year ranged from 81.9% to 96.4% except in 2015 when it was 63.7% (Table 1). The number of ADAP clients captured in our analytic dataset ranged from 224,110 in 2015 to a peak of 256,004 in 2020, then declined to 227,955 in 2022. The number of ADAP clients who were included in the study for each year ranged from 146,879 (65.5%) in 2015 to 220,839 (96.9%) in 2022. Jurisdictions excluded each year (and reasons for exclusion) are detailed in Supplemental Table 1.

**Table 1.** Total Number and Percentage of People with HIV (PWH), AIDS Drug Assistance Program (ADAP) Clients, and Non-ADAP Clients Included for Each Year of Study, 2015-2022.

| **Year** | **Total PWH in United States**<br>**n** | **PWH Included in Study**<br>**n (row %)** | **ADAP Clients in United States**<br>**n** | **ADAP Clients Included in the Study**<br>**n (row %)** | **Non-ADAP Clients in United States**<br>**n** | **non-ADAP Clients Included in the Study**<br>**n (row %)** |
| --- | --- | --- | --- | --- | --- | --- |
| 2015 | 942,988 | 601,102 (63.7) | 224,110 | 146,879 (65.5) | 718,878 | 454,223 ( 63.2 ) |
| 2016 | 967,557 | 841,798 (87.0) | 222,830 | 194,191 (87.1) | 744,727 | 647,607 ( 87.0 ) |
| 2017 | 991,448 | 857,051 (86.4) | 230,068 | 199,903 (86.9) | 761,380 | 657,148 ( 86.3 ) |
| 2018 | 1,014,481 | 887,817 (87.5) | 234,798 | 205,257 (87.4) | 779,683 | 682,560 ( 87.5 ) |
| 2019 | 1,036,801 | 898,406 (86.7) | 248,811 | 212,264 (85.3) | 787,990 | 686,142 ( 87.1 ) |
| 2020 | 1,049,709 | 860,176 (81.9) | 256,004 | 205,836 (80.4) | 793,705 | 654,340 ( 82.4 ) |
| 2021 | 1,068,119 | 911,114 (85.3) | 242,985 | 212,487 (87.4) | 825,134 | 698,627 ( 84.7 ) |
| 2022 | 1,092,023 | 1,052,239 (96.4) | 227,955 | 220,839 (96.9) | 864,068 | 831,400 ( 96.2 ) |
| 2015-2022 | 1,020,391 | 863,713 ( 84.6 ) | 235,945 | 199,707 ( 84.6 ) | 784,446 | 664,006 ( 84.6 ) |

In all years analyzed, >90% of individuals were eligible for VS assessment. ADAP clients consistently had lower VL test coverage than overall PWH population in the same year (Table 2). Among PWH included, 96.5-97.4% had a reported VL test each year; 97.0-99.5% of non-ADAP clients and 90.2-96.3% of ADAP clients had a reported test.

**Table 2.** Total Number and Percentage of People with HIV (PWH), AIDS Drug Assistance Program (ADAP) Clients, and non-ADAP Clients who were Eligible for Viral Suppression Assessment, who had Viral Suppression (VS), and and who had Detectable Viral Load (VL), 2015-2022.

| Year |  | 2015 | 2016 | 2017 | 2018 | 2019 | 2020 | 2021 | 2022 | 2015-2022 |
| --- | --- | --- | --- | --- | --- | --- | --- | --- | --- | --- |
| **People with HIV (PWH)** |  |  |  |  |  |  |  |  |  |  |
| Total Number in Study | n | 601,102 | 841,798 | 857,051 | 887,817 | 898,406 | 860,176 | 911,114 | 1,052,239 | 863,713 |
| Eligible for VS assessment<sup>1</sup> | n (%) | 581,886 (96.8) | 820,091 (97.4) | 830,295 (96.9) | 862,045 (97.1) | 871,670 (97.0) | 838,191 (97.4) | 882,317 (96.8) | 1,015,556 (96.5) | 837,756(97.0) |
| VS<sup>2</sup> | n (%) | 349,076 (60.0) | 504,441 (61.5) | 524,349 (63.2) | 558,413 (64.8) | 572,529 (65.7) | 544,011 (64.9) | 584,798 (66.3) | 661,758 (65.2) | 537,422 (64.2) |
| Detectable VL<sup>3</sup> | n (%) | 232,810 (40.0) | 315,650 (38.5) | 305,946 (36.8) | 303,632 (35.2) | 299,141 (34.3) | 294,180 (35.1) | 297,519 (33.7) | 353,798 (34.8) | 300,334 (35.8) |
| **AIDS Drug Assistance Program (ADAP) Clients** |  |  |  |  |  |  |  |  |  |  |
| Total Number in Study | n | 146,879 | 194,191 | 199,903 | 205,257 | 212,264 | 205,836 | 212,487 | 220,839 | 199,707 |
| Eligible for VS assessment<sup>1</sup> | n (%) | 141,467 (96.3) | 186,245 (95.9) | 180,389 (90.2) | 186,649 (90.9) | 198,728 (93.6) | 194,210 (94.4) | 196,077 (92.3) | 206,445 (93.5) | 186,276 (93.3) |
| VS<sup>2</sup> | n (%) | 114,877 (81.2) | 162,114 (87.0) | 157,655 (87.4) | 168,003 (90.0) | 180,234 (90.7) | 177,443 (91.4) | 176,847 (90.2) | 186,268 (90.2) | 165,430 (88.8) |
| Detectable VL<sup>3</sup> | n (%) | 26,590 (18.8) | 24,131 (13.0) | 22,734 (12.6) | 18,647 (10.0) | 18,494 (9.3) | 16,767 (8.6) | 19,230 (9.8) | 20,177 (9.8) | 20,846 (11.2) |
| **Non-AIDS Drug Assistance Program (ADAP) Clients** |  |  |  |  |  |  |  |  |  |  |
| Total Number in Study | n | 454,223 | 647,607 | 657,148 | 682,560 | 686,142 | 654,340 | 698,627 | 831,400 | 664,006 |
| Eligible for VS assessment<sup>1</sup> | n (%) | 440,419 (97.0) | 633,846 (97.9) | 649,906 (98.9) | 675,396 (99.0) | 672,942 (98.1) | 643,981 (98.4) | 686,240 (98.2) | 809,111 (97.3) | 651,480 (98.1) |
| VS<sup>2</sup> | n (%) | 234,199 (53.2) | 342,327 (54.0) | 366,694 (56.4) | 390,410 (57.8) | 392,295 (58.3) | 366,568 (56.9) | 407,951 (59.4) | 475,490 (58.8) | 371,992 (57.1) |
| Detectable VL<sup>3</sup> | n (%) | 206,220 (46.8) | 291,519 (46.0) | 283,212 (43.6) | 284,986 (42.2) | 280,647 (41.7) | 277,413 (43.1) | 278,289 (40.6) | 333,621 (41.2) | 279,488 (42.9) |
| **Chi-square<sup>4</sup>** |  |  |  |  |  |  |  |  |  |  |
| Eligible for VS assessment<sup>1</sup> | p | < 0.0007 | < 0.0007 | < 0.0007 | < 0.0007 | < 0.0007 | < 0.0007 | < 0.0007 | < 0.0007 | < 0.0007 |
| VS<sup>2</sup> | p | < 0.0007 | < 0.0007 | < 0.0007 | < 0.0007 | < 0.0007 | < 0.0007 | < 0.0007 | < 0.0007 | < 0.0007 |
| Detectable VL<sup>3</sup> | p | < 0.0007 | < 0.0007 | < 0.0007 | < 0.0007 | < 0.0007 | < 0.0007 | < 0.0007 | < 0.0007 | < 0.0007 |

1. Eligible for VS assessment: All PWH and non-ADAP clients were eligible for the calendar year's viral suppression assessment if the person is diagnosed by the end of the previous calendar year and alive at the end of the calendar year of the calculation; an ADAP client is eligible for the calendar year's assessment of viral suppression if the person had a reported viral load test during that calendar year
2. VS: Of those eligible for VS assessment, the number who had VS (< 200 copies/mL)
3. Detectable Viral Load: Of those eligible for VS assessment, the number who had a detectable VL (> 200 copies/mL)
4. Chi-square test comparing between ADAP Clients and Non-ADAP clients

VS for PWH ranged from 60-66.3% while VS for non-ADAP clients ranged from 53.2-59.4% and for ADAP clients, it ranged from 81.2%-91.4% (Table 2). In five of the eight years, the VS rates for ADAP clients was > 90.0%. The VS rate among PWH increased from 2015-2019, declined in 2020, increased in 2021, and decreased in 2022. The VS rate among ADAP clients increased from 2015-2020, decreased in 2021, and plateaued in 2022.

In all years, the proportion of PWH with VS who were ADAP clients was greater than the proportion of PWH who were ADAP clients (Figure 1). ADAP clients were overrepresented among those with VS. Over 2015-2022, the proportion of PWH who were ADAP clients was 23.1%, and the proportion of PWH with VS who were ADAP clients was 30.8% (Figure 1).

**Figure 1.**
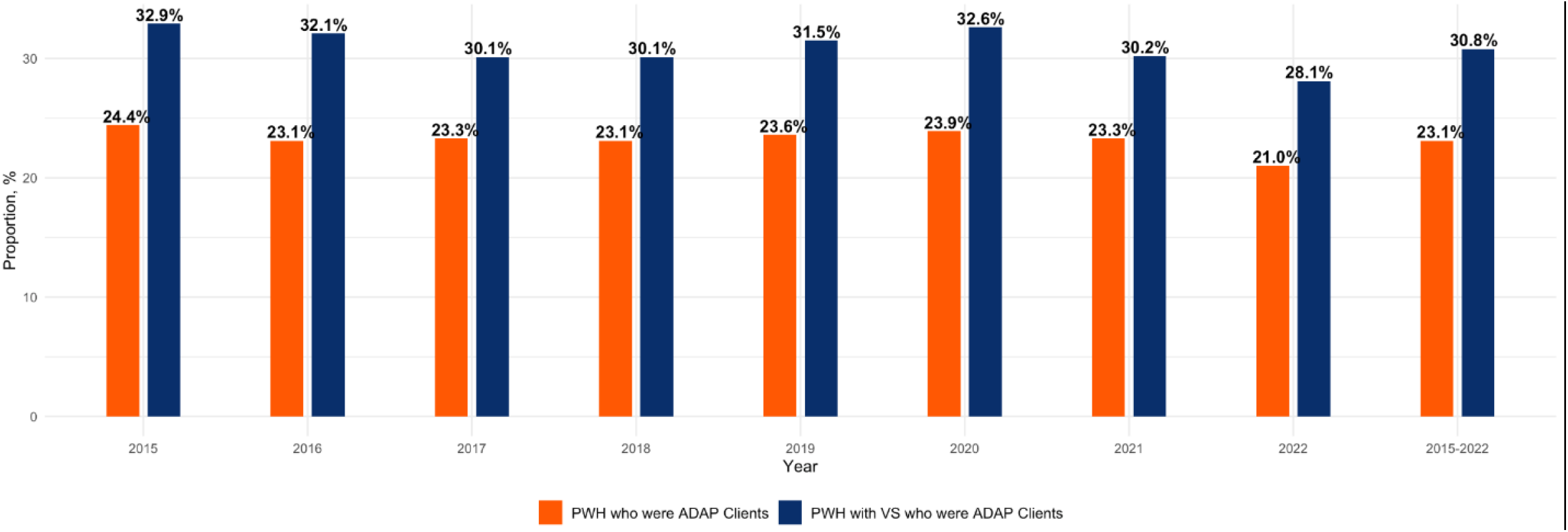
Proportion of People with HIV (PWH) who were ADAP Clients and Proportion of PWH with Viral Suppression (VS) who were ADAP Clients, 2015-2022

In all years, the proportion of PWH who were ADAP clients was greater than the proportion of PWH who were eligible for VS assessment who were ADAP clients (Supplemental Figure 1). ADAP clients were underrepresented among those eligible for VS assessment. For 2015-2022, while the proportion of PWH who were ADAP clients was 23.1%, the proportion of PWH who were eligible for VS assessment who were ADAP clients was 22.2% (Supplemental Figure 1).

In all years, the proportion of PWH who were ADAP clients was greater than the proportion of PWH with detectable VLs who were ADAP clients (Supplemental Figure 2). ADAP clients were underrepresented among those with detectable VLs. In 2015, 11.4% of PWH with detectable VLs were ADAP clients. The lowest proportion of PWH with detectable VLs who were ADAP clients was in 2020 and 2022 at 5.7% despite ADAP clients being 23.9% and 21.0% of all PWH during those years. For 2015-2022, while the proportion of PWH who were ADAP clients was 23.1%, the proportion of PWH with detectable VLs who were ADAP clients was 7.1% (Supplemental Figure 2).

### Sensitivity Analyses

The findings were robust under all three missing-data assumptions. In the *worst-case scenario*, where all individuals with unknown VL were assumed to be *not* virally suppressed, overall VS rates declined slightly for both groups. However, ADAP clients continued to demonstrate considerably higher VS rates than non-ADAP clients (Supplemental Table 2). For example, in 2020, the national VS dropped from 64.9% (as observed) to 63.2% under this conservative assumption, while the ADAP client VS rate dropped from 91.4% to 86.2%. Although attenuated, the VS rate for ADAP clients remained markedly higher than that of non-ADAP clients across all years and scenarios. Estimates from both the *base-case*, where all individuals with unknown VL were assumed to have the observed rate of VS, and the *best-case*, where unknown VL was treated as VS, also consistently demonstrated higher VS rates among ADAP clients compared to non-ADAP clients.

Notably, across all scenarios, ADAP clients consistently comprised approximately one-third of all virally suppressed PWH nationwide, despite only account for 21.0% to 24.4% of the total PWH (Supplemental Figure 3). Similarly, sensitivity analyses for detectable VL yielded no qualitative changes (Supplemental Table 3); under the most extreme assumptions, ADAP clients consistently comprised only a small minority of those with unsuppressed VLs each year (Supplemental Figure 4).

### Comparative Statistics

In formal comparisons, ADAP clients differed significantly from non-ADAP clients on all key indicators. ADAP clients were slightly less likely to be counted in the VS eligibility cohort (reflecting their lower VL test coverage) yet significantly more likely to achieve VS than non-ADAP individuals. Even under the most conservative missing-data assumption (worst-case scenario), ADAP clients’ VS over non-ADAP clients remained statistically significant. Likewise, ADAP clients had a significantly lower proportion with detectable VL compared to non-ADAP clients in each year. All differences reported were significant at <0.0007.

## Discussion

The role that ADAP plays to ensure that PWH reach VS – a key metric for the effectiveness of HIV interventions – is profound. A CDC modeling study demonstrated that helping PWH achieve VS is one of the two most cost-effective ways to reduce new HIV infections and end the HIV epidemic.^16^ Over our study period, almost a third of the entire VS rate was due to ADAP clients having VS, despite ADAP serving less than a quarter of PWH. In other words, the fact that ADAP clients are more likely to be virally suppressed as compared to the broader population of PWH is responsible for a disproportionate percentage of the total national VS rate. This finding is substantial, and our sensitivity analyses—including scenarios in which all unknown VLs were assumed to be detectable—suggest that ADAP’s contribution to the national VS rate is not simply an artifact of differential VL reporting between ADAP and non-ADAP clients.

ADAP clients being overrepresented among those with VS and underrepresented among those with detectable VLs demonstrates that ADAPs are helping ADAP clients, a vulnerable and often marginalized population with lower incomes, to achieve good outcomes. With sustained VS, the lifespan of PWH approaches that of people without HIV.^17^ Sustained VS also results in lower HIV-related morbidity and mortality, maintains or improves CD4 cell counts, and prevents HIV drug resistance.^18–21^ Having less ADAP clients who have detectable VLs also contributes to community health by reducing community HIV VLs and subsequently reducing HIV transmission.^22,23^ A CDC study estimated that 63% of new HIV infections were transmitted from the 34% of PWH who knew about their HIV diagnosis and had a detectable VL.^24^

The ability of ADAP to contribute so substantially to VS is even more noteworthy given the program’s relatively flat federal funding over the past decade.^25^ As ADAP enrollment has continued to increase and as list prices of HIV medications have continued to rise, the annual federal allocation for ADAP has not increased.^5,26^ Yet, ADAPs are helping some of the most vulnerable PWH achieve high rates of VS and are making an outsized contribution to the US’ VS.

National and state partners working to end HIV in the US should examine how to better integrate ADAP into state and local programs, including opportunities to maximize ADAP enrollment, streamline ADAP re-certifications, and increase access to ADAP-subsidized health insurance. Additional efforts should ensure that ADAP maximizes uptake among subgroups of PWH across age, gender, race/ethnicity, and other sociodemographic factors. This is essential given that enrollment in ADAP has been associated with increased engagement in care for women with HIV,^27^ ADAP-supported health insurance has been associated with increased VS,^28–30^ and dis-enrollment from ADAP has been associated with loss of VS.^31^

Further investing in ADAP, through EHE funds or other federal and state funds, may have significant gains in VS.^6^ Currently, ADAP does not directly receive any EHE funding. Instead, the majority of funding goes directly to city and county governmental public health programs. The EHE funding allocated for RWHAP recipients goes to RWHAP Part A and RWHAP Part B recipients, and while there is nothing prohibiting recipients from allocating EHE funding to ADAP, these allocations have been largely absent from EHE programming.^8^

Finally, federal policymakers should pay more attention to the role that ADAP plays in leveraging manufacturer discounts for HIV medications. The ability of ADAP to manage year-on-year utilization increases with flat federal funding is likely largely dependent on its access to 340B discounts as well as additional price concessions negotiated directly with manufacturers. As newer products (with higher price tags) make their way through research, development, and FDA approval, additional examination of ADAP’s ability to facilitate access to those products for un and under-insured low-income PWH will be critical. Policymakers should also pay attention to the cost-effectiveness of ADAP insurance assistance compared to direct provision of medications and the potential impact that increasing insurance enrollment for ADAP clients has on VS rates.^32^ Possible cuts to domestic HIV programs, including the EHE initiative, being considered by Congress and the Administration could jeopardize the success of ADAPs in supporting VS.

We found that ADAP clients were less likely to be eligible for VS assessment in a given year than non-ADAP clients. This may be due to their VLs not being reported to the state at all or not being reported in a timely manner. It is possible that ADAP clients, PWH with lower incomes, could be facing additional barriers to having a VL test in a given year compared non-ADAP clients.

Some strengths of this analysis include the longitudinal nature of the data and the combination of ADAP-specific data with HIV surveillance data to give a better picture of ADAP’s role in the HIV healthcare delivery system in the US. This study has several limitations. First, the higher VS rate among ADAP clients could also be impacted by the other non-ADAP RWHAP services these clients may be receiving, something not captured in the data. One study that assessed the role of receipt of RWHAP clinical and support services in helping PWH to achieve VS indicated that PWH with low-incomes were more likely to achieve VS if they received care at a RWHAP-funded facility.^33^ More analysis is needed to understand the interplay between ADAP and other RWHAP services on HIV outcomes. Second, the VS measure is a snapshot aggregate rate and may not indicate client-level sustained VS over time. And third, even though the overall study participation is high, the geographic areas that were excluded from the study because of missing or inaccurate data are more likely to be areas with low HIV prevalence, which may limit application of our findings nationally and for EHE.

ADAPs play a critical role in ending the HIV epidemic in this country. Federal investment in ADAP sustainability and innovation combined with state and local analysis of replicable ADAP best practices could help to increase VS rates and ultimately slow or end new HIV transmissions.

## Data Availability

All data used are publicly available online through NASTAD's National RWHAP Part B and ADAP Monitoring Project Annual Reports and the the Centers of Disease Control and Prevention's (CDC) National Center for HIV/AIDS, Viral Hepatitis, STD, and TB Prevention (NCHHSTP) AtlasPlus.

https://nastad.org/adap-monitoring-project

https://www.cdc.gov/nchhstp/about/atlasplus.html

## Article Information

## Acknowledgments

The authors would like to thank state and territorial Ryan White HIV/AIDS Program Part B and AIDS Drug Assistance Program (ADAP) teams for their time and effort in completing the National RWHAP Part B Monitoring Survey, and NASTAD for their time and effort in conducting the ADAP annual survey and writing the annual report. The authors would like to thank the Centers of Disease Control and Prevention’s (CDC) National Center for HIV/AIDS, Viral Hepatitis, STD, and TB Prevention (NCHHSTP) for their time and effort on AtlasPlus.

## Author Contributions

All authors contributed substantively to this manuscript in the following ways: conception and design of the study (K.A.M., A.K, T.H., E.T.R.M.), acquisition of data (K.A.M, F.L.), data analysis (K.A.M., E.R., F.L.), interpretation of results (all authors), manuscript drafting (K.A.M., A.K., E.R.), editing and revisions (all authors), and final approval of submitted version (all authors).

## Funding

This work was supported by the National Institute of Allergy and Infectious Diseases at the National Institutes of Health (Grant No. R01AI170093 to K.A.M.). The content is solely the responsibility of the authors and does not necessarily represent the official views of the National Institutes of Health.

## Potential Conflicts of Interest

All authors have completed and submitted the International Committee of Medical Journal Editors form for disclosure of potential conflicts of interest. During the conduct of this study and the writing of the manuscript, all authors report receiving support from the National Institute of Allergy and Infectious Diseases with payments made to their respective institutions. Unrelated to the submitted work, the following disclosures were reported from the past 36 months: K.A.M. also reports unpaid leadership position: Chair of the Advisory Committee to Virginia Medication Assistance Program. A.S. reports stock ownership: Merck & Company and Organon stock. A.K. reports being a paid consultant for NASTAD and JSI.

