## Supplemental Tables and Figures for "State AIDS Drug Assistance Programs’ Contribution to the United States’ Viral Suppression, 2015-2022"

*Supplemental Tables*

Supplemental Table 1…………………………………………………………………………………..………2

Supplemental Table 2………………………………………………………………………………………..…3

Supplemental Table 3……………………………………………………………………………………….….4

*Supplemental Figures*

Supplemental Figure 1…………………………………………………………………………………….……5

Supplemental Figure 2…………………………………………………………………………………….……6

Supplemental Figure 3…………………………………………………………………………………….……7

Supplemental Figure 4…………………………………………………………………………………….……8

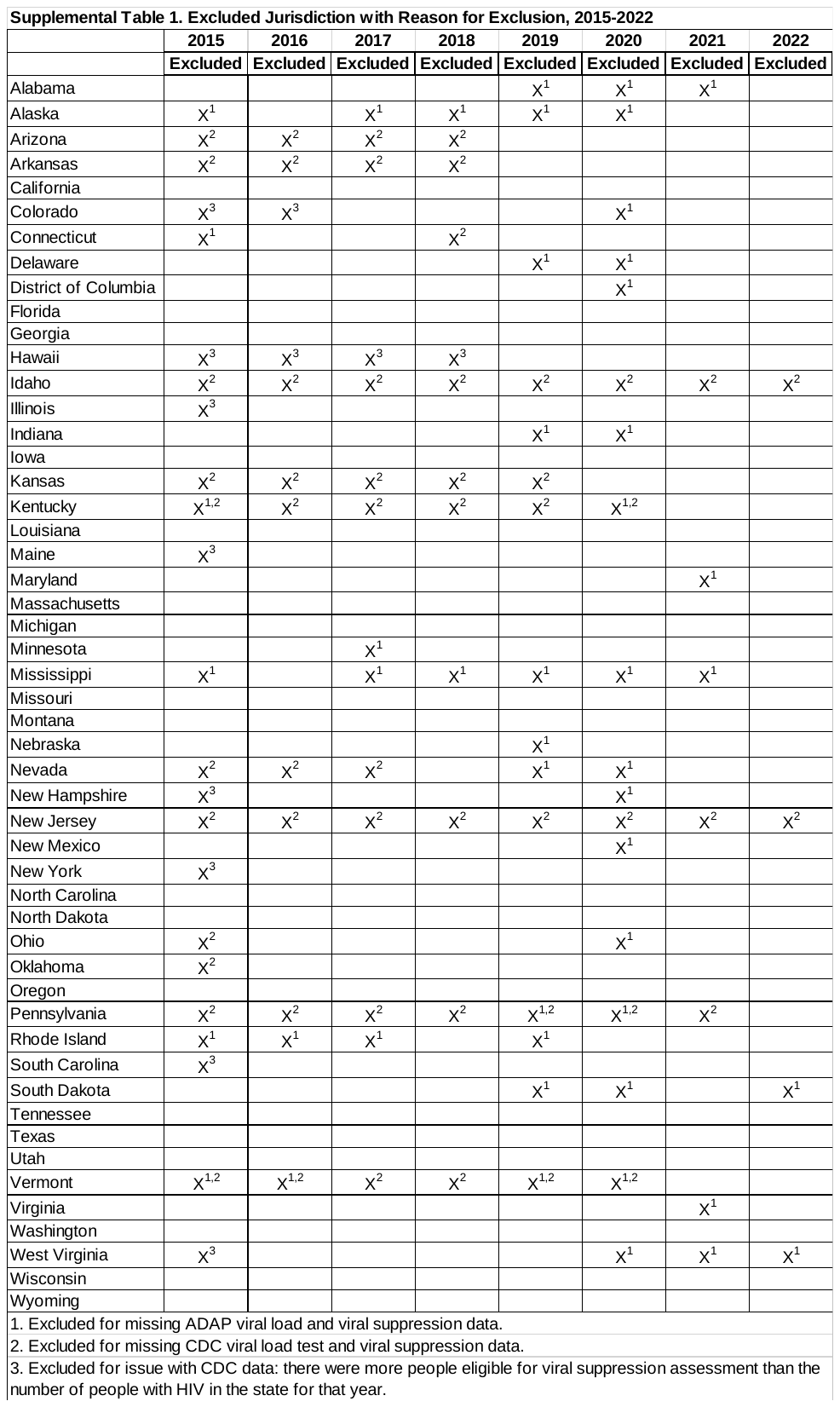

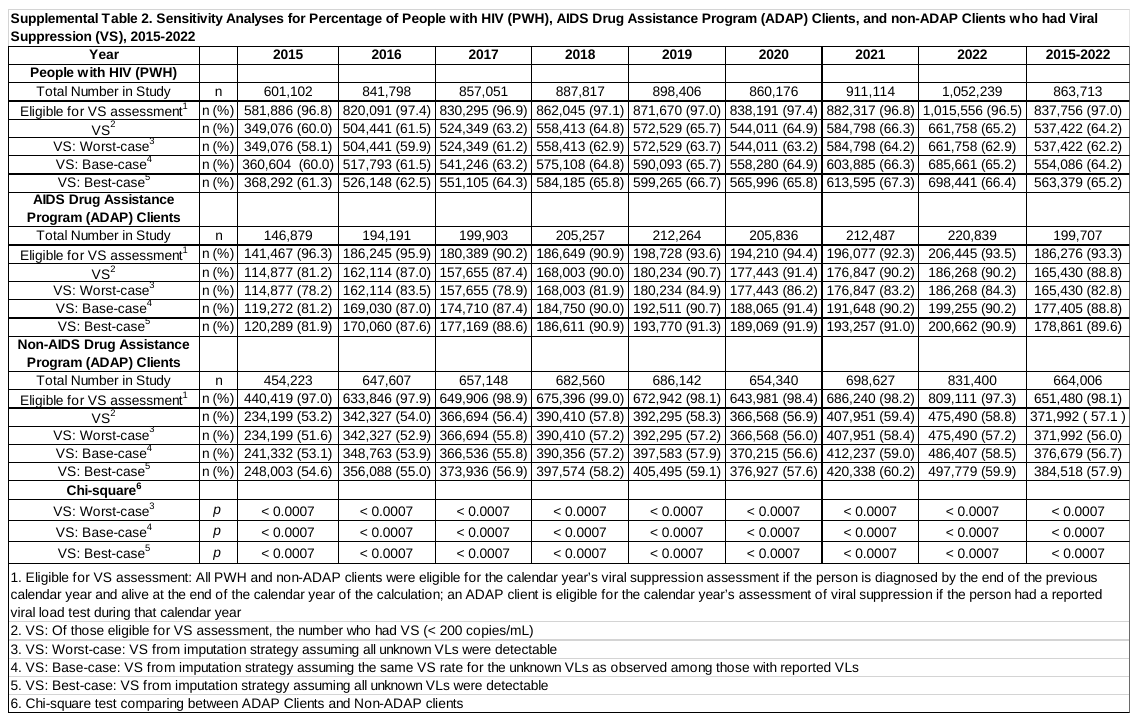

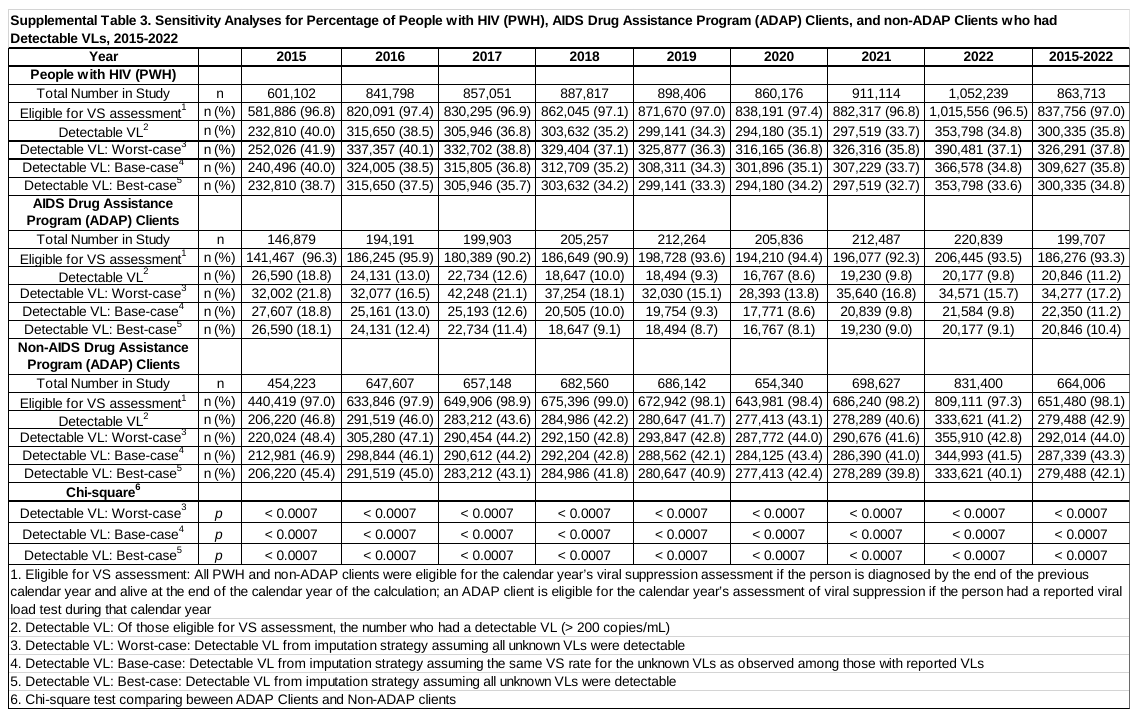

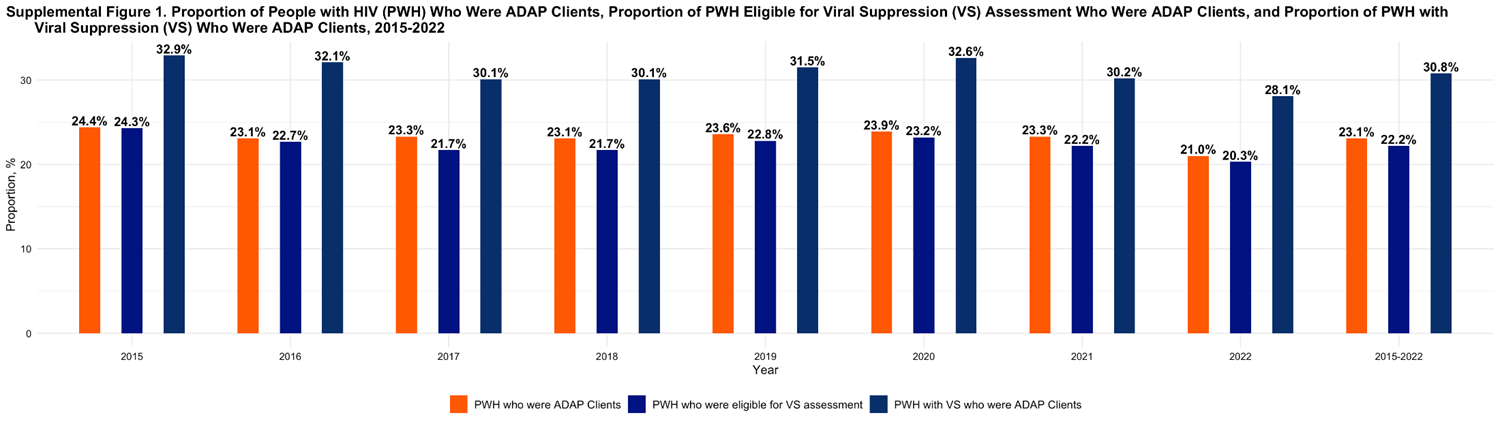

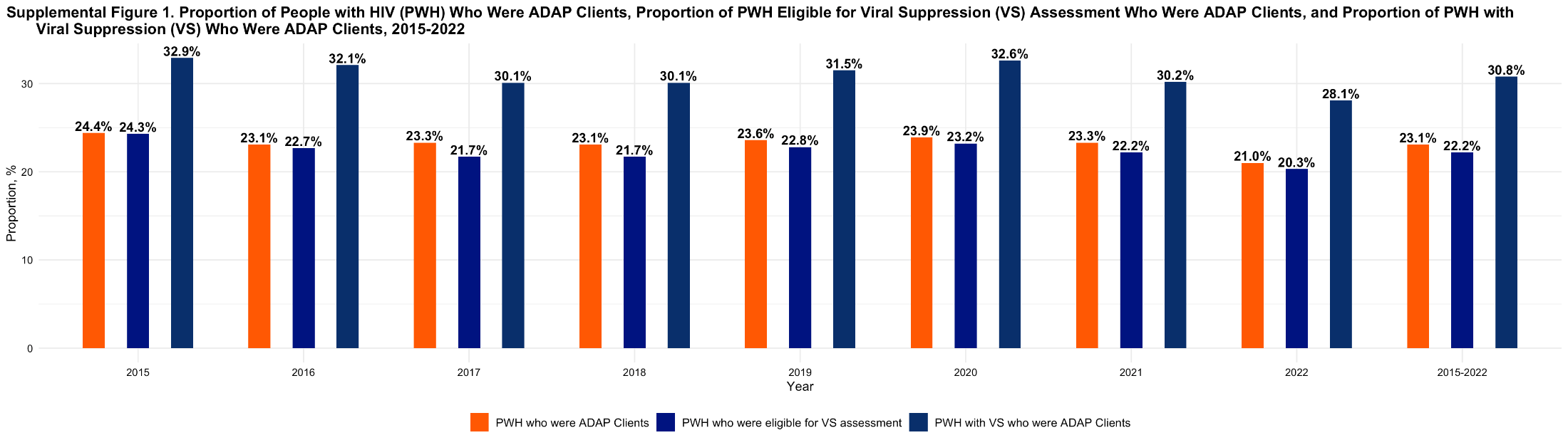

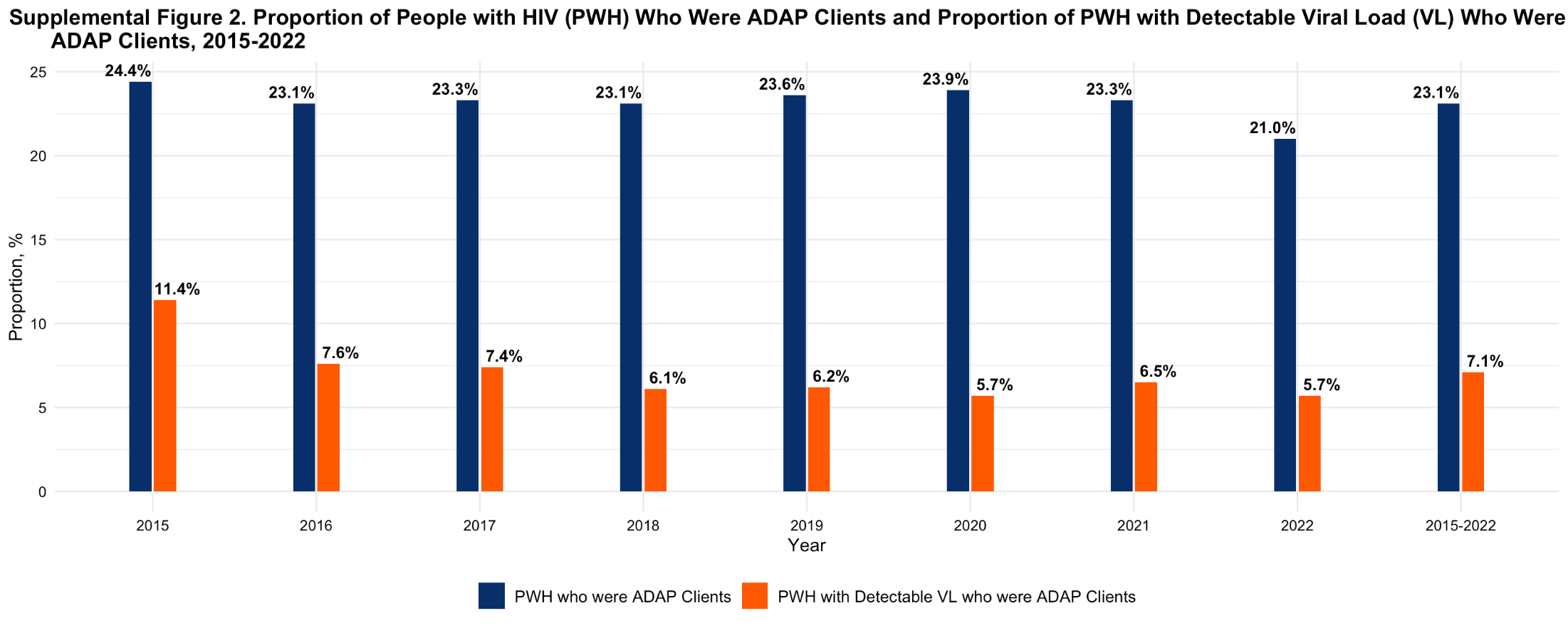

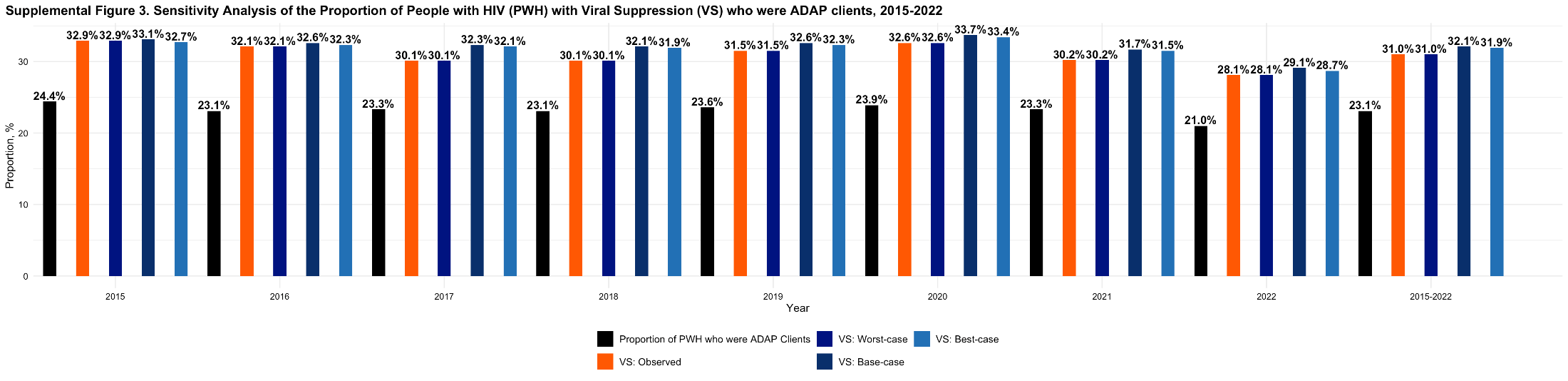

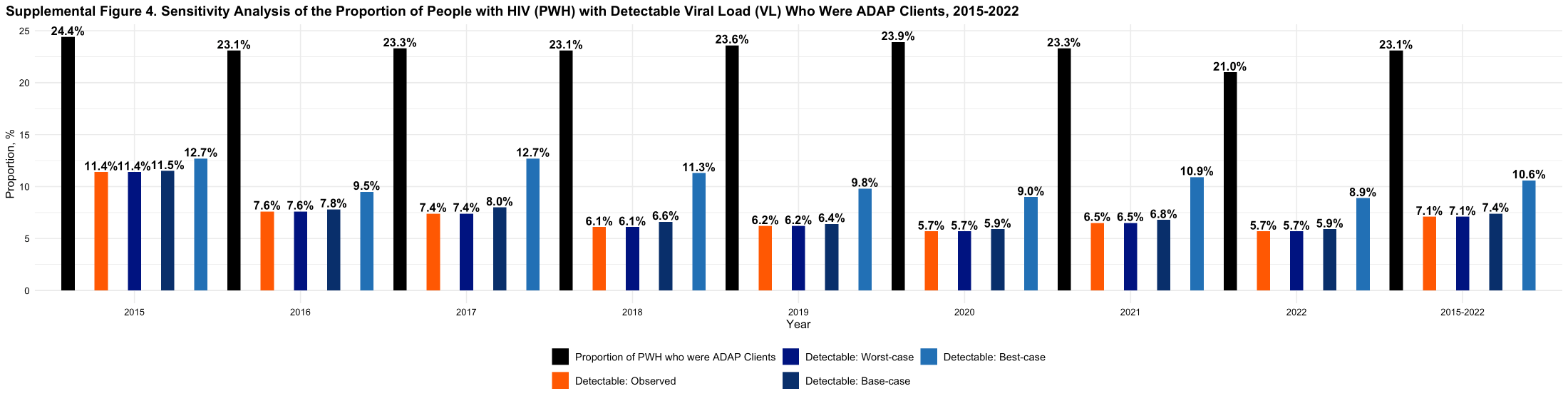
